# Hospital admissions in inflammatory rheumatic diseases during the COVID-19 pandemic: incidence and role of disease modifying agents

**DOI:** 10.1101/2020.05.21.20108696

**Authors:** Benjamín Fernandez-Gutierrez, Leticia Leon, Alfredo Madrid, Luis Rodriguez-Rodriguez, Dalifer Freites, Judit Font, Arkaitz Mucientes, Jose Ignacio Colomer, Juan Angel Jover, Lydia Abasolo

## Abstract

**Background:** In this pandemic, it is essential for rheumatologist and patients to know the relationship between COVID-19 and inflammatory rheumatic diseases (IRD). We want to assess the role of targeted synthetic or biologic disease modifying antirheumatic drugs (ts/bDMARDs) and other variables in the development of moderate-severe COVID-19 disease in IRD.

**Methods:** An observational longitudinal study was conducted (1^st^Mar to 15^th^Apr 2020). All patients from the rheumatology outpatient clinic from a hospital in Madrid with a medical diagnosis of IRD were included. Main outcome: hospital admission related to COVID-19. Independent variable: ts/bDMARDs. Covariates: sociodemographic, comorbidities, type of IRD diagnosis, glucocorticoids, NSAIDs and conventional synthetic DMARDs (csDMARDs). Incidence rate (IR) of hospital admission related to COVID-19, was expressed per 1,000 patients-month. Cox multivariate regression analysis was run to examine the influence of ts/bDMARDs and other covariates on IR.

**Results:** 3,591 IRD patients were included (5,896 patients-month). Concerning csDMARDs, methotrexate was the most used followed by antimalarial. 802 patients were on ts/bDMARDs, mainly anti-TNF agents, and rituximab. Hospital admissions related to COVID-19 occurred in 54 patients (1.36%) with an IR of 9.15 [95%CI: 7-11.9]. In the multivariate analysis, older, male gender, presence of comorbidities and specific systemic autoimmune conditions (Sjoegren, polychondritis, Raynaud and mixed connective tissue disease) had more risk of hospital admissions regardless other factors. Exposition to ts/bDMARDs did not achieve statistical signification. Use of glucocorticoids, NSAIDs, and csDMARDs dropped from the final model.

**Conclusion:** This study provides additional evidence in IRD patients regarding susceptibility to moderate-severe infection related to COVID-19.

## INTRODUCTION

New SARS-CoV-2 causes a myriad of clinical signs and symptoms with analytic typical features. As a whole, all characteristics are called COVID-19 disease,^1^ and it has affected millions of lives worldwide.

A majority of COVID -19 patients present no symptoms or mild symptomatology. Other smaller subgroup, shows progression to a moderate disease. A further subgroup apparently develops a syndrome with autoimmunity and/or autoinflammatory features with critical/fatal outcomes ^2,3^. In this sense, it seems that COVID-19 disease is having a particular incidence and severity in patients with advanced age and comorbidities, mainly diabetes, hypertension, ischemic heart disease, and previous respiratory diseases ^4,5^.

Serious infection is a well-recognized cause of morbidity and mortality across a number of inflammatory rheumatic diseases (IRD). In this context of pandemic, it is essential for rheumatologist and for patients themselves to know the relationship between COVID-19 and IRD. Several guidance’s for the management of inflammatory rheumatic patients in this scenario, based in expert opinion, have been performed, ^6–8^ as there is scarce epidemiological research of the potential risk of IRD and/or disease modifying antirheumatic drugs (DMARDs) on COVID-19 disease and its severity. Few experiences from Italy and Spain have been recently published, showing that patients with chronic inflammatory arthritis treated with biologic or synthetic DMARDs do not seem to be at increased risk of infection nor respiratory complications from SARS-CoV-2 compared with the general population ^9–11^. These preliminary findings, if corroborated, could be very relevant and helpful for the IRD patient management. The purpose of this study is to estimate the incidence rate of moderate-severe COVID-19 disease, globally and stratified by age, sex, type of diagnosis and therapy used in IRD patients from our health area. Then, we want to assess the role of exposition to targeted synthetic or biologic DMARDs (ts/bDMARDs) in the development of moderate-severe COVID-19 disease, taking into account all other relevant parameters, such as age, sex, comorbidity, conventional synthetic DMARDs (csDMARDs), corticosteroids, NSAIDs and type of rheumatic diagnosis.

## METHODS

### Setting, design, and patients

The setting is a tertiary hospital of the Public Health System of the Community of Madrid, the Hospital Clínico San Carlos (HCSC), covering a catchment area of 400,000 people.

We performed a retrospective observational study, from the 1^st^ of March 2020, (when our health area had the first hospital admission related to COVID-19) to the 15^th^ of April 2020. All patients being attended at the rheumatology outpatient clinic of HCSC, whose data were recorded in the health clinical record of our service (HRC Penelope), and with at least one visit in the previous year, were preselected. We included all patients >16 years old, with medical diagnosis (according to ICD-10) of inflammatory rheumatic disease including: **a)** chronic inflammatory arthritis: rheumatoid arthritis (RA), psoriatic arthritis (PSA), spondyloarthritis (SPA), uveitis, Inflammatory bowel disease, Juvenile idiopathic arthritis, and inflammatory polyarthritis (IA); **b)** systemic autoimmune mixed connective tissue disease conditions: Sjögren’s syndrome (Sjo), systemic sclerosis, mixed connective tissue disease (MCTD); systemic lupus erythematosus (SLE), polymyalgia rheumatic (PMR), vasculitis, Behcet’s syndrome, sarcoidosis, polychondritis, autoinflammatory syndrome, antiphospholipid syndrome, inflammatory myopathies and, primary Raynaud phenomenon. Those with loss of follow-up at the time of inclusion were excluded.

The study was conducted in accordance with the Declaration of Helsinki and Good Clinical Practices and was approved by the HCSC institutional ethics committee (approval number 20/268-E_BS).

### Variables

The primary outcome was the development of moderate-severe COVID-19 disease defined as hospital admission related to COVID-19 during the study period. This definition was based on medical diagnosis +/− PCR diagnostic test. The independent variable was exposure to ts/bDMARDs incluidng: a) anti-TNF alfa (Infliximab, adalimumab, etanercept, certolizumab, golimumab); b) Other Biologics: anti-IL6 (tocilizumab, sarilumab); rituximab (Rtx); abatacept (Abata); belimumab (Beli); anti-IL17 / 23; anti-IL17 (ustekunumab, ixekizumab, secukinumab); c) Jakinibs (JAKi: tofacitinib, baricitinib).

As covariables we considered: **1)** Sociodemographic baseline characteristics including sex, age and IRD duration. **2)** Type of IRD: including chronic inflammatory arthritis and systemic autoimmune conditions. **3)** Baseline comorbidity described in table 1.**4)** Other chronic treatment for IRD: a) glucocorticoids, b) NSAIDs, c) csDMARDs incluidng: leflunomide (Lef); methotrexate (Mtx); azathioprine or mycophenolate mophetilo (Aza), cyclophosphamide; cyclosporine (Cpa); d) other csDMARDs inlcuidng: antimalarial (Am: chloroquine / hydroxychloroquine); sulfasalazine (Ssz); and colchicine.

**Table 1.** Baseline demographic and clinical characteristics among IRD patients

| **Variable** | **All IRD patients**<br>**n =3,951** |
| --- | --- |
| Women, n (%) | 2,857 (72.3) |
| Age, mean (SD), years | 61.8 (16.6) |
| Disease evolution time, mean (SD), years | 10.80(8.38) |
| Smoking habit (Active) | 170 (4.3) |
| Diagnosis, n (%) |  |
| Rheumatoid arthritis | 1,486 (37.7) |
| Inflammatory Poliarthritis | 170 (4.3) |
| Axial spondyloarthritis | 491 (12.4) |
| Psoriatic arthritis | 289 (7.3) |
| Polymyalgia rheumatic | 377 (9.5) |
| Systemic lupus erythematosus | 248 (6.3) |
| Mixed connective tissue disease | 158 (4.0) |
| Systemic sclerosis | 80 (2.0) |
| Sjogren's syndrome | 146 (3.7) |
| Vasculitis | 115 (2.9) |
| Behcet disease | 43 (1.1) |
| Polychondritis | 16 (0.6) |
| Polymyositis | 35 (0.89) |
| Raynaud | 92 (2.3) |
| Uveitis | 100 (2.5) |
| Others* | 104 (2.6) |
| Comorbidities, n (%) |  |
| Hypertension | 860 (21.8) |
| Dyslipidemia | 707 (17.9) |
| Depression | 250 (6.3) |
| Diabetes Mellitus | 323 (8.2) |
| Heart disease** | 296 (7.5) |
| Ischemic vascular disease*** | 181 (4.6) |
| Chronic liver disease | 127 (3.2) |
| Chronic kidney disease | 57 (1.5) |
| Lung disease (ILD/COPD) | 312 (7.9) |
| History or presence of cancer | 235(5.9) |
| Venous Thrombosis/lung embolism | 54 (1.4) |
| Thyroid disease | 430 (10.9) |
| NSAIDs use, n (%) | 860 (21.7) |
| Glucocorticoid use, n (%) | 1,804 (45.6) |
| Colchicine use, n(%) | 56(1.4) |
| csDMARDs, n (%): |  |
| Mtx-Lef-Aza | 1,961 (49.6) |
| Cpa | 27(0.68) |
| Ssz | 317 (8.0) |
| Am | 666 (16.8) |
| ts/bDMARDs, n (%) |  |
| Anti-TNF | 521 (13.2) |
| Ifx | 52 (1.3) |
| Ada | 188 (4.7) |
| Etn | 117(2.9) |
| Certo | 103(2.6) |
| Goli | 61(1.5) |
| Non Anti-TNF | 246 (6.2) |
| Abata | 27(0.68) |
| Tozi | 42(1.06) |
| Rtx | 122(3.1) |
| Sari, Secu, lxe, Uste | 49 (1.2) |
| Beli | 6(0.15) |
| JAKi , n (%) | 35 (0.89) |
| Bari | 27(0.68) |
| Tofa | 8(0.2) |

**Abbreviations:** IRD: inflammatory rheumatic disease; SD: standard deviation; IQR: Interquartile range; COPD: chronic obstructive pulmonary disease; ILD: interstitial lung disease. Others\*: Inflammatory bowel disease, antiphospholipid syndrome, juvenile idiopathic arthritis, autoinflammatory syndromes; sarcoidosis. \*\*Heart disease: arrhythmias, valvulopathies, cardiomyopathies, heart failure. Ischemic vascular disease: stroke, cardiovascular and peripheral vascular disease. csDMARD: Conventional synthetic disease-modifying anti rheumatic drug. ts/bDMARDs target synthetic/biologic disease-modifying anti rheumatic drug; Anti-TNF: Tumor necrosis factor-alpha inhibitor (infliximab (Ifx), adalimumab (Ada), etanercept (Etn), certolizumab (Certo), golimumab (Goli). Other bios includes: rituximab (Rtx), abatacept (Abata), tocilizumab (Tozi), sarilumab (Sari), secukinumab (Secu), ustekinumab (Uste), ixekizumab (Ixe) and belimumab (Beli). JAKi: JAK inhibitors including (tofacitinib, baricitinib); csDMARD: Conventional synthetic disease-modifying anti rheumatic drug.

To consider patients were exposed to drugs, treatment had to start at least one month before the beginning of the study, had to continue during the study period until the end of study or medical admission for Am, glucocorticoids, Ssz, and NAIDs. Regarding Mtx, Lef, Aza, Cpa and ts/bDMARDs, treatment had to start at least one month before the beginning of the study, had to continue during at least 21st of March, end of study or hospital admission. In the case of Rtx, the last infusion had to be at least on January.

### Data sources

Patient sociodemographic, clinical, and therapeutic data were obtained through the HCR Penelope. SARS-CoV-2 PCR diagnostic tests information were obtained from the microbiology service of HCSC (n=5,577 patients with PCR test in the study period). Central Services of the Hospital provided us all the HCSC admissions (n=1,146 in the study period). All information from IRD patients was merged.

### Statistical Analysis

Patient’s characteristics were described as mean and standard deviation for continuous variables, while proportions are shown for categorical variables.

Survival techniques were used to estimate the incidence rate of hospital admissions related to CVID-19 (IR). IR was given per 1,000 persons-months with a 95% confidence interval [CI]. The time of observation comprised the elapsed time from 1st March 2020, to the date of patient’s hospital admission, or end of study.

The incidence rate ratio of hospital admissions related to COVID-19 among IRD patients and population from our heath area older than 16 years was assessed.

Cox bivariate analyses were done in IRD to evaluate statistical differences between hospital admission risks and all variables. Cox multivariate regression model (adjusted by age, sex, type of diagnosis and comorbidities) was run to examine the possible influence of ts/bDMARDs in hospital admissions regardless other factors. In the model we also included glucocorticoids, csDMARDs, and all other variables with a p<0.2 from the bivariate analysis. Results were expressed as hazard ratio (HR) and [CI]. Proportional hazard assumption was tested using the scaled Schoenfeld residuals. A two-tailed *p* value under 0.05 was considered to indicate statistical significance.

## RESULTS

### IRD Patients description

3,951 IRD patients were included, with a total follow up of 5,896 patients-months. As we shoe in table 1, many of them were women in their sixties. The most frequent diagnosis was RA, followed by SPA, PMR, PSA and SLE. Regarding comorbidities, hypertension, dyslipemia, thyroid disease and diabetes mellitus were the most prevalent. Concerning csDMARDs, Mtx was the most used (n=1,461), followed by Am, Lef (n=333) Slz, and Aza (n=245). 6 patients were using cyclophosphamide. 32% of the patients did not use csDMARDs, 47% were on monotherapy and the remaining 21% used at least two concomitant csDMARDs (mainly Mtx+Am; Mtx+Ssz and Mtx+Lef). Concerning ts/bDMARD (n= 802), 12.5% of them were on monotherapy and the remaining 87.5% combined with csDMARDs. The most frequent were anti-TNF agents, followed by Rtx.

Hospital admissions related to COVID-19 occurred in 54 patients (1.36%) during the follow-up. 76% were positive to PCR test, 5% were negative and in the remaining 19% the PCR test was not performed.

### Incidence rate of hospital admission related to COVID-19

The IR was estimated in 9.15 [7-11.9] per 1,000 patients-months. As expected, IR has been increasing throughout the study: when we analyzed in fortnightly cuts, the IR from March 1 to March 15 was 1.01 per 1,000; for March 15 to March 30 was 6.3 per 1,000; and for April 1 to April 15 was 6.6 per 1.000 patients. In fact, IR in the period of March 15 to April 15 was higher, estimated in 13 per 1.000 patients.

For IRD, the cumulative incidence of hospital admissions related to COVID-19 during the study period was 15 per 1,000 patients, whereas the cumulative incidence for hospitalized patients related to COVID-19 (n=1,059) in our health area (n>16 years: 325.900)^12^ was lower being estimated in 3.2 per 1,000 persons (incidence rate ratio: 4.6 [3.4-6.1]; p=0.000).

As shown in table 2 the crude IR could vary depending on different variables. It was higher for men than for women and in those older compared to youngers. It seemed lower for those included in the chronic inflammatory arthritis group compared to those from the systemic autoimmune conditions, with the exception of SLE. It was similar in patients with or without csDMARDs. None hospital admissions were found for patients with Cpa, colchicine, nor cyclophosphamide. Finally, concerning ts/bDMARDs IR was higher in patients on Rtx and lower in patients using anti-TNF. None hospital admissions were found for patients with Abata, Sari, Secu, Uste, Ixe nor Beli.

**Table 2:**
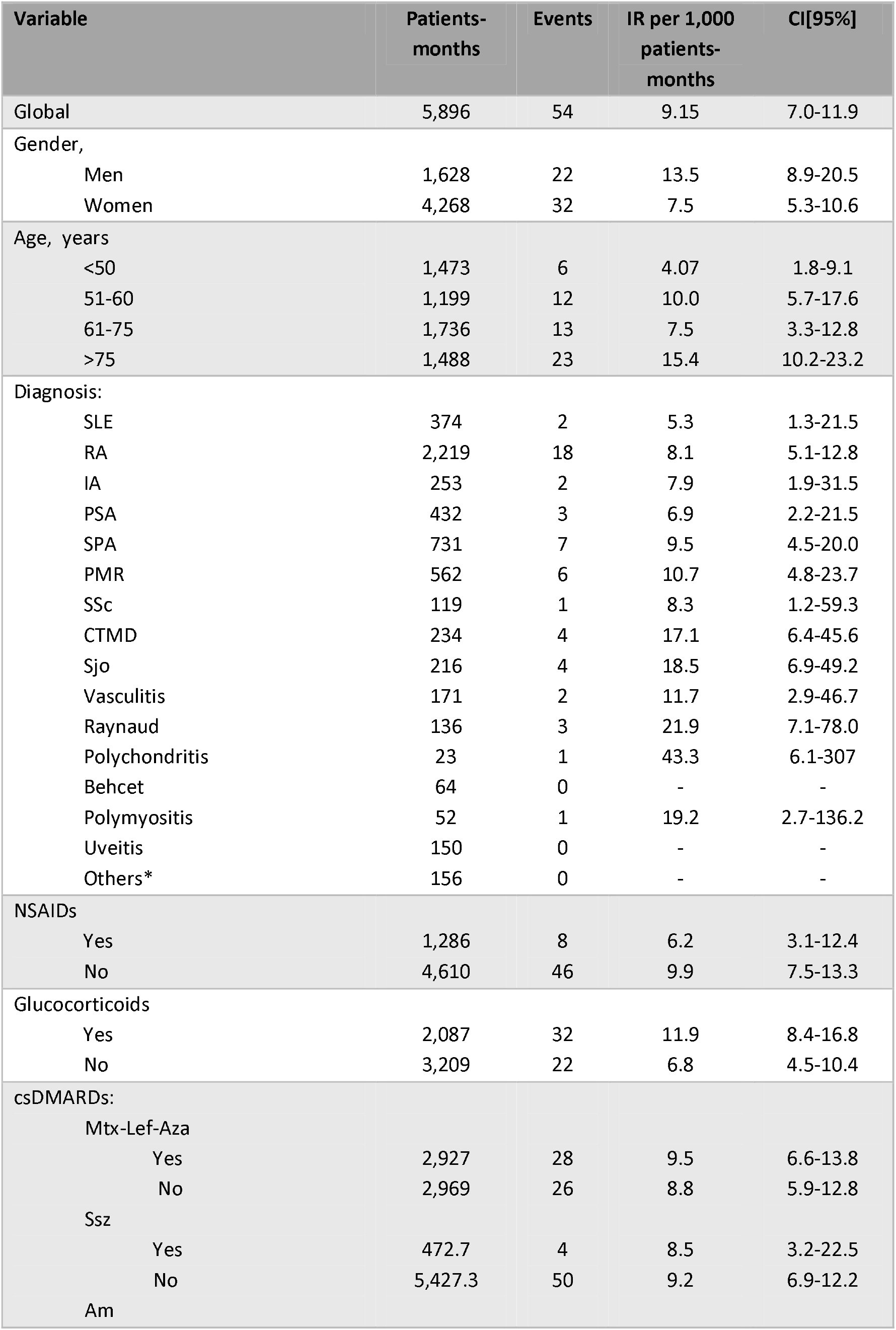
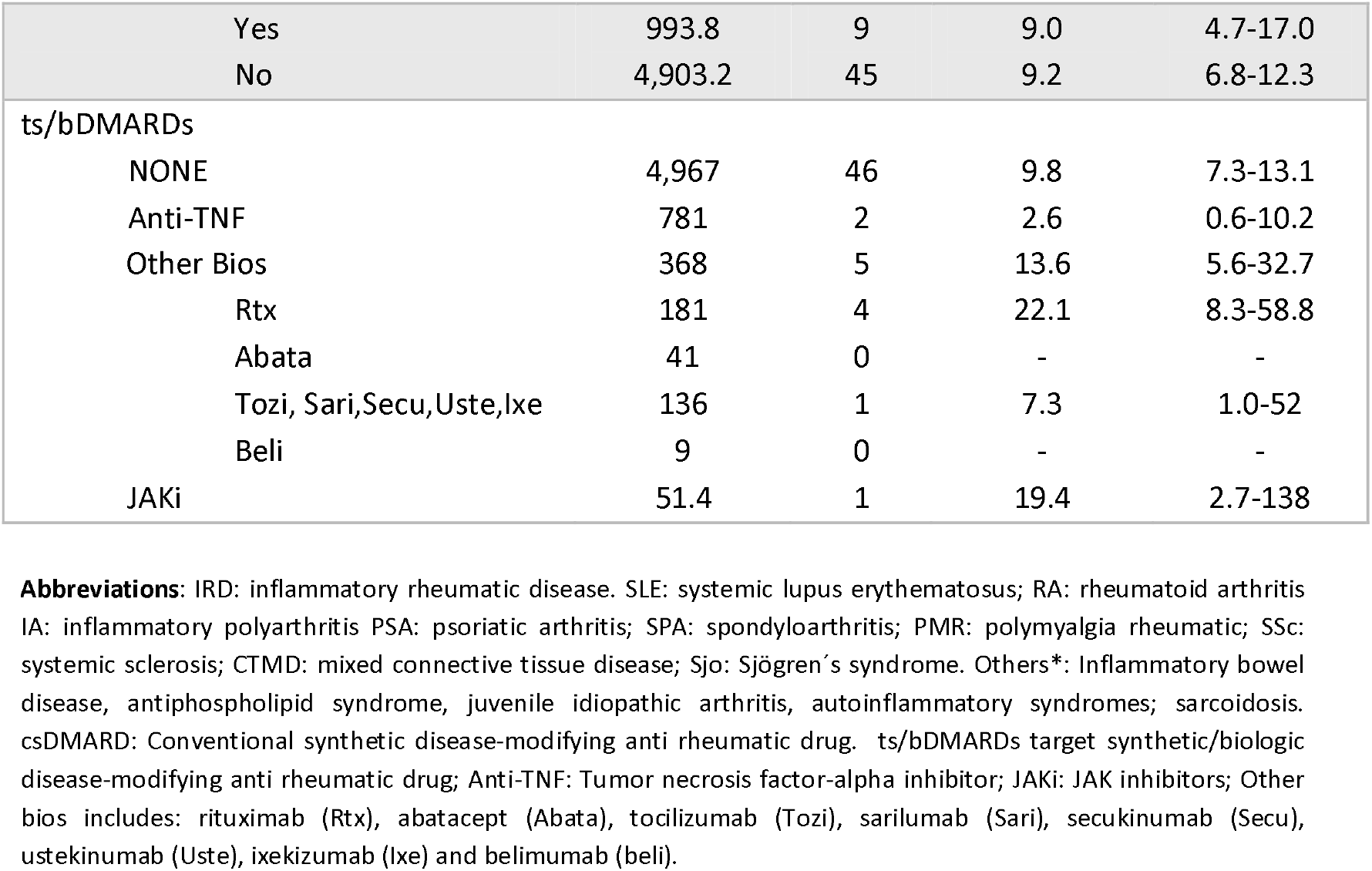
Incidence rate of hospital admissions related to COVID 19 in IRD patients

### Bivariate analysis

As expected age, sex and several comorbidities, but also the use of glucocorticoids was statistical associated to hospital admission related to COVID-19 in IRD. NSAIDs and ts/bDMARDs did not achieve statistical significance, but had a trend (table 3). When we analyzed separately other biologics, Rtx compared with the rest, had a trend of more risk of hospital admission (HR: 2.2 [0.85-2.4], p=0.1). Regarding type of diagnosis, some systemic autoimmune conditions had a trend of more risk of hospital admission except for SLE that had lower risk (p=0.4). SLE versus chronic inflammatory arthritis did not reach statistical significance (HR: 0.68 [0.16-2.8], p=0.59). However, other systemic autoimmune conditions (not SLE) versus chronic inflammatory arthritis, achieved a trend of more risk (HR: 1.62 [0.94-2.8], p=0.08).

**Table 3.**
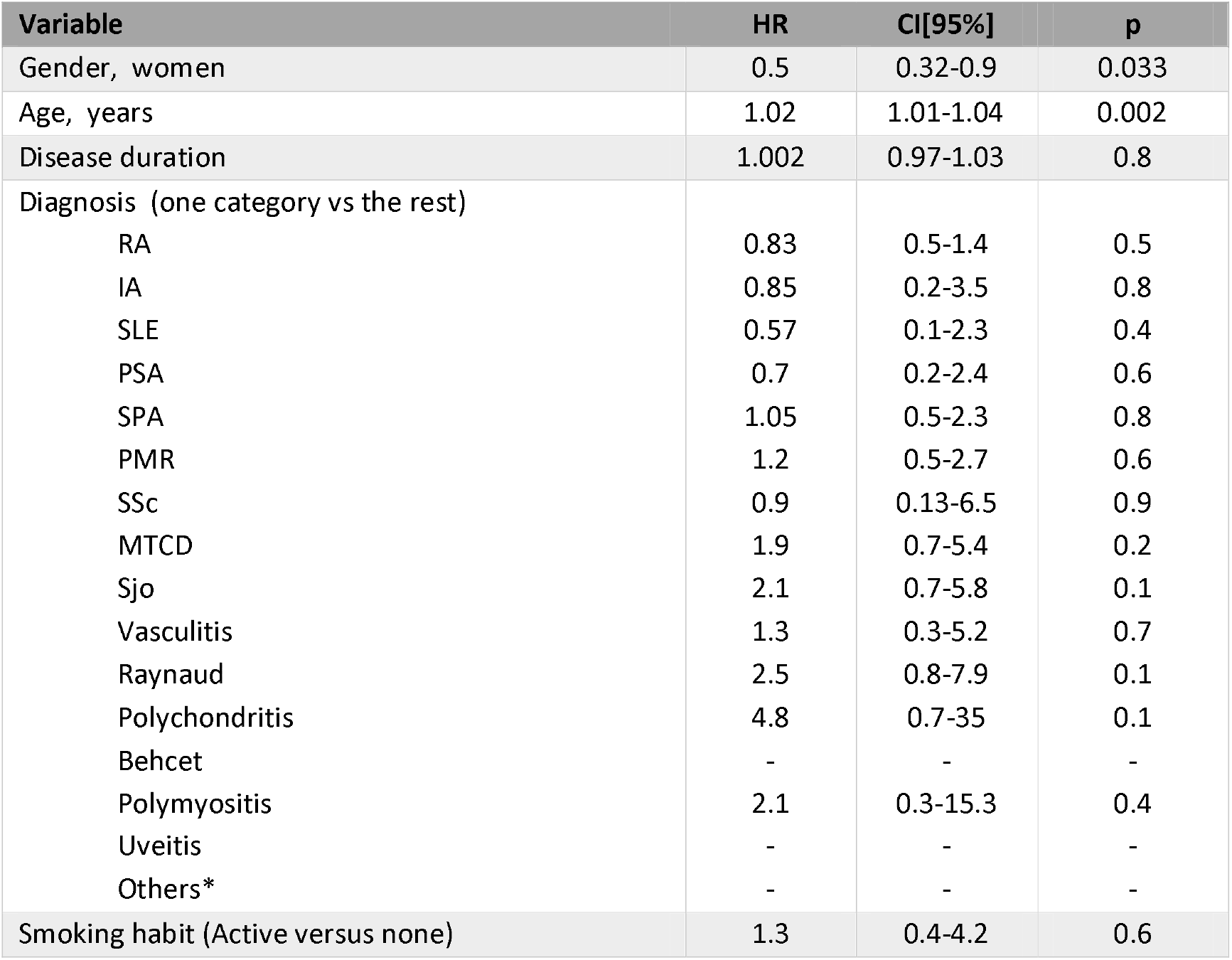
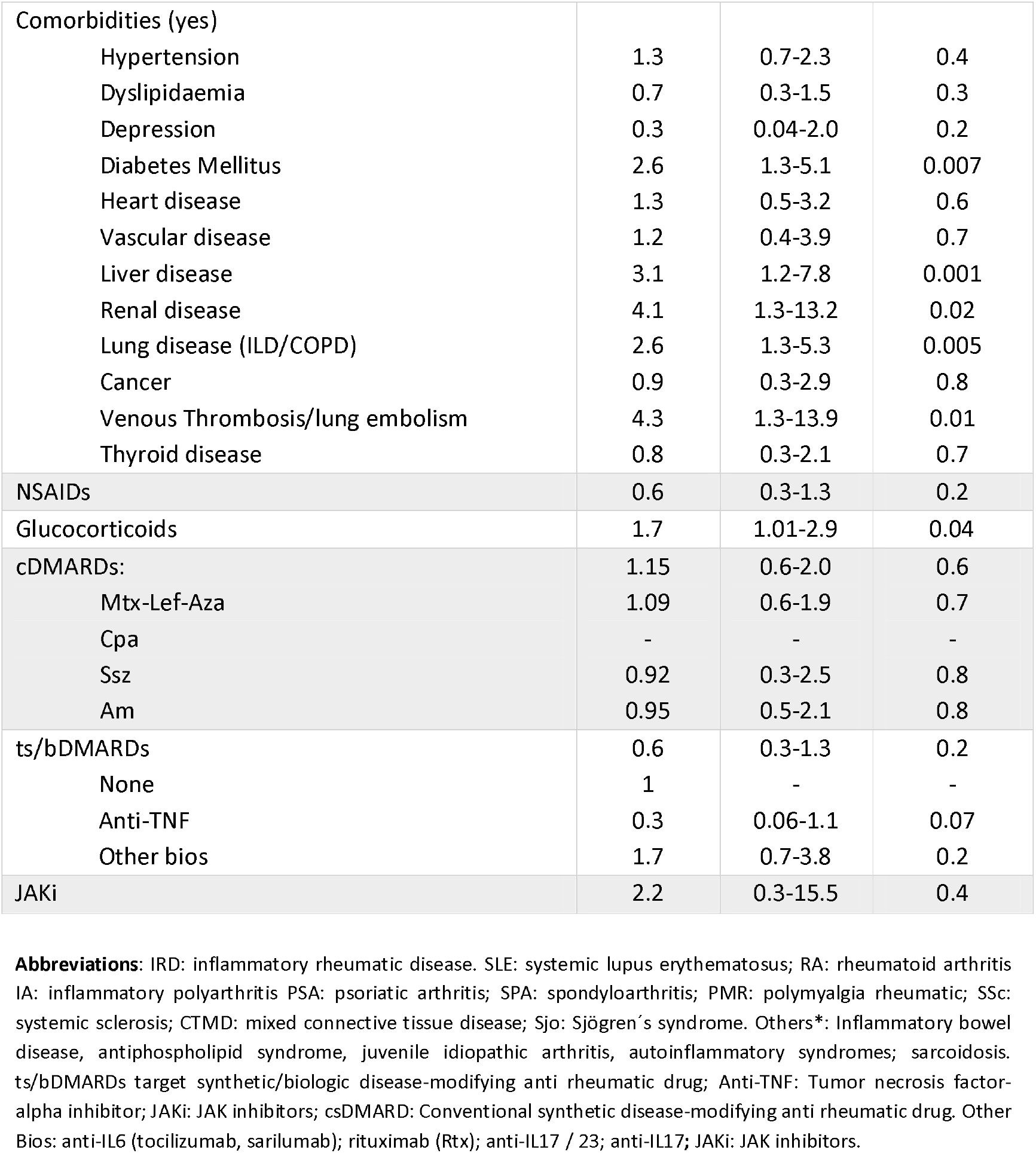
Hazard ratio of medical admission related to COVID-19 in IRD patients. Bivariate analysis

### Multivariate analysis

In the final model, after adjusting by sex, age, comorbidities and type of diagnosis, ts/bDMARDs did not achieve statistical significance compared to none use (table 4). Regarding specific non-TNFs vs none, they did not reach statistical significance either (Rtx HR: 2[0.71-5.6] p=0.190; Jakis HR: 2.6[0.3-19.3] p=0.3; and Tozi HR: 2.2[0.3-16.3] p=0.4).

**Table 4.** Role of ts/bDMARDs on risk of hospital admission related to COVID-19 in IRD patients. Adjusted by rheumatic diagnosis, age, sex, csDMARDs and comorbidity. Multivariate analysis

| Variable | HR | CI[95%] | p |
| --- | --- | --- | --- |
| Gender, women | 0.54 | 0.3-0.94 | 0.03 |
| Age, years |  |  |  |
| <50 | 1 | - | - |
| 51-74 | 1.84 | 0.75-4.5 | 0.18 |
| >75 | 1.59 | 1.3-6.5 | 0.04 |
| Diagnosis: Systemic autoimmune conditions vs chronic inflammatory arthritis | 1.27 | 0.7-2.23 | 0.3 |
| Comorbidities (yes) | 2.18 | 1.2-3.8 | 0.03 |
| ts/bDMARDs |  |  |  |
| None | 1 | - | - |
| Anti-TNF | 0.32 | 0.07-1.37 | 0.125 |
| Non Anti-TNF | 1.58 | 0.67-3.8 | 0.290 |

**Abbreviations:** IRD: inflammatory rheumatic disease. Systemic autoimmune conditions (PMR; SSC, Sjo, CTMD, Vasculitis, Raynaud, Polymyositis, Polychondritis; Behcet, sarcoidosis, antiphospholipid syndrome, SLE) vs chronic Inflammatory arthritis (RA; IA; JIA, PSA, Spa, Uvetis, Inflammatory bowel disease). Comorbidities including the presence of at least one of the follows: ischemic vascular disease, diabetes mellitus, venous thrombosis/lung embolism, chronic kidney disease, liver disease, lung disease (ILD/COPD). ts/bDMARDs target synthetic/biologic disease-modifying anti rheumatic drug; Anti-TNF: Tumor necrosis factor-alpha inhibitor; Non Anti-TNF: anti-IL6 (tocilizumab, sarilu mab); rituximab (Rtx); anti-IL17 / 23; anti-IL17+JAKi: JAK inhibitors.

Interestingly, glucocorticoids (HR: 1.46[0.8-2.5]; p=0.18), Am (HR: 1.3[0.6-2.8]; p=0.46), Ssz (HR: 1.24[0.4-3.6]; p=0.6), Mtx-Lef-Aza (HR: 1.25[0.7-2.2]; p=0.4), and NAIDs (HR: 0.9[0.4-2.2]; p=8), dropped from the final model.

Concerning diagnosis, systemic autoimmune conditions vs chronic inflammatory arthritis did not achieved statistical significance. When we categorized this variable in specific diagnoses (we grouped RA-PSA as reference category based on syndromic similarity and incidence rates), the final model did not change but we found some interesting results: Sjoegren (HR: 3[1.01-9.03]; p=0.04), Raynaud (HR: 4[1.2-13.7]; p=0.02), and polychondritis (HR: 111[1.5-91]; p=0.02) increased the risk of hospital admission compared to RA-PSA and independently of other factors. SLE (HR: 0.99[0.2-4.3]; p=0.8) did not achieve statistical significance. MCTD (HR: 2.5 [0.8-7.4]; p=0.09) achieved a trend or more risk, and the HR in rest of diagnosis did not differ (p>0.2).

When analyzing the final model using as variables specific comorbidities instead of the presence of comorbidities, lung disease (HR: 2.1[1.03-4.2], p=0.03), liver disease (HR: 3.1 [1.2-8.0], p=0.01), and venous thrombosis/lung embolism (HR: 3.4[1.1-11.3], p=0.03) achieved statistical significance.

The proportionality of these regression models was tested with a p value=0.7.

## DISCUSSION

In this real-world longitudinal study of 1.5 months, we include a broad spectrum of inflamatory rheumatic diseases treated with or without ts/bDMARDs, csDMARDs, and glucocorticoids. With all this information, we have been able to estimate the incidence rate of hospital admissions related to COVID-19 in ILD, but also to evaluate the influence of ts/bDMARDs, csDMARDs, types of IRD, and other factors in the risk of hospital admissions related to COVID-19.

This pandemic has had a great impact, especially in Madrid, with more than 41,304 hospital admissions until first week on May ^13^. In this study we have been able to show the rising of this incidence from March to April.

In our study, the IR of hospital admissions related to COVID-19 in IRD patients was estimated in 9.15 per 1,000 patients-months. When we compare the IR of hospital admissions related to COVID-19 among IRD patients and the reference population, it seems that IRD have an increased risk. Age, sex, therapies, and disease specific factors contribute form sure. Other studies have compared the IR of IRD with their reference population without differences^10,11,14^. But they have compared PCR confirmed cases regardless the severity. Otherwise, two of them ^10,11^ did not include patients with systemic autoimmune conditions. Moreover, the incidence rate varies per regions and time period ^10, 11, 14–16^.

Regarding ts/bDMARDs, the crude IR of hospital admission related to COVID-19 found in our study was lower for those on Anti-TNF and higher for those with non-TNF biologics. But in the multivariate analysis the sligthly statistical differences from the bivariate analysis dissapeared. Interestingly only one hospital admission related to COVID-19 was found on tocilizumab, and none were found on abatacept, anti IL-17/23 nor baricitinib. This may be promising, but we should also bear in mind that the number of patients on these drugs were not sufficient to draw specific conclusions. But, in agreement with other authors ^15–17^ ts/bDMARDs and mainly Anti-TNF, do not seem to be associated with worse outcomes in IRD.

Another interesting finding of this paper, is that the crude IR of hospital admissions related to COVID-19 differs among rheumatologic diseases, being somewhat higher in the systemic autoinmune conditions. In the multivariate analysis, these differences remained statistically significant for Sjoegren, polychondritis, CTMD, but also for primary Raynaud. Nevertheless, LES had the same risk as RA-PSA without statistical significance. Other systemic autoimmune conditions did not reach statistical significance, but maybe the number of patients was not enough to find those differences.

Regarding other therapies, the crude incidence rates seems to be similar in patients with and without csDMARDs, higher in those on corticoids, and lower in those using NSAIDs. Nevertheless, after the multivariate analysis none of them remained statistically significant. According to Favalli et al ^16^, it seems that Mtx, Lef or Aza do not increase the risk of hospital admission related to COVID-19. In the case of Am, several autors have published its beneficial effect for the acute treatment of moderate-severe infection related to COVID-19 ^18,19^ In agreement of other authors ^20,21^ we are not able to demonstrate the protective effect of the chronic use of Am on moderate-severe infection related to COVID-19. Regarding glucocorticoids, although the crude incidence rate was higher, they dropped from the final model. Nevertheless, these results should be corroborated analyzing corticoids by doses.

Interestingly, we corroborated the role of age, male gender and comorbidities ^2,3^ in the susceptibility to develop moderate-severe COVID-19 disease. Specifically liver disease, lung disease and venous thrombosis/lung embolism achieved statistical significance in the multivariate analysis. Ischemic vascular disease and diabetes mellitus only a trend and hypertension, cancer or dyslipidemia did not achieved statistical signification. We do not have to forget that data was recorded during routine consultations, with a heavy workload environment making easier the possibility of incomplete information mainly related to comorbidity.

It is true that the PCR test should be required as a part of the main outcome definition. However, in all admissions included, almost 20% of them did not have the PCR performed due to a lack of available tests and or extreme health care overload at that time. Nevertheless, all were reviewed being clinically compatible and managed as COVID-19. But, if we exclude these cases, the real incidence of hospital admissions related to COVID-19 would be underestimated. Another limitation is that we could have lost hospital admissions that have gone to other hospitals. Two of them were rescued for analysis, and we think there won’t be many more considering the state of alarm and confinement decreed in Spain since March 14. As strengths, we include 3,591 non-selected patients with a board spectrum of IRD, with not standardized immunosuppressive therapy reflecting clinical practice from our health area, being able to adjust for confounders.

To our knowledge, this is the largest study to date outlining the severity of COVID-19 in terms of hospital admissions in IRD. It seems that patients with IRD could have a higher susceptibility of moderate-severe COVID-19 disease compared to the general population, maybe at expenses of systemic autoimmune diseases rather than chronic inflammatory arthritis. Moreover, we have been able to analyze in greater extent the safety surrounding the administration of disease modifying treatments. It seems that predisposition to develop moderate-severe COVID-19 disease in IRD, is at expenses of the type of diagnosis, age, sex and comorbidities, rather than the treatments exposed including ts/bDMARDs and csDMARDs.

## Data Availability

Data will be available upon request

## Acknowledgements

The authors would also like to thank Ana M Perez for their help in the data collection. A special thank you to all rheumatologist and nurses colleagues who contributed in the care of the patients in an innovative and so involved way.

## Contributors

BF, LL, JAJ, LRR and LA contributed to the conception and design of the study. DF, JF, AMG, AM, JIC and LL were involved in data collection. LA, AMG and JIC were involved in data base management. LA and LL performed the data analysis and interpretation of data. All authors contributed to drafting and/or revising the manuscript.

## Ethics approval

The study was approved by the Hospital Clínico San Carlos institutional ethics committee (approval number 20/268-E-BS). This study was conducted according to the principles of the Declaration of Helsinki.

## Patient and Public Involvement

Patients or the public were not involved in the design, or conduct, or reporting, or dissemination plans of our research.

